# Investigation of Autosegmentation Techniques on T2-Weighted MRI for Off-line Dose Reconstruction in MR-Linac Adapt to Position Workflow for Head and Neck Cancers

**DOI:** 10.1101/2021.09.30.21264327

**Authors:** Brigid A. McDonald, Carlos Cardenas, Nicolette O’Connell, Sara Ahmed, Mohamed A. Naser, Kareem A. Wahid, Jiaofeng Xu, Daniel Thill, Raed Zuhour, Shane Mesko, Alexander Augustyn, Samantha M. Buszek, Stephen Grant, Bhavana V. Chapman, Alexander Bagley, Renjie He, Abdallah Mohamed, John P. Christodouleas, Kristy K. Brock, Clifton D. Fuller

## Abstract

**Purpose:** In order to accurately accumulate delivered dose for head and neck cancer patients treated with the Adapt to Position workflow on the 1.5T magnetic resonance imaging (MRI)-linear accelerator (MR-linac), the low-resolution T2-weighted MRIs used for daily setup must be segmented to enable reconstruction of the delivered dose at each fraction. In this study, our goal is to evaluate various autosegmentation methods for head and neck organs at risk (OARs) on on-board setup MRIs from the MR-linac for off-line reconstruction of delivered dose.

**Methods:** Seven OARs (parotid glands, submandibular glands, mandible, spinal cord, and brainstem) were contoured on 43 images by seven observers each. Ground truth contours were generated using a simultaneous truth and performance level estimation (STAPLE) algorithm. 20 autosegmentation methods were evaluated in ADMIRE: 1-9) atlas-based autosegmentation using a population atlas library (PAL) of 5/10/15 patients with STAPLE, patch fusion (PF), random forest (RF) for label fusion; 10-19) autosegmentation using images from a patient’s 1-4 prior fractions (individualized patient prior (IPP)) using STAPLE/PF/RF; 20) deep learning (DL) (3D ResUNet trained on 43 ground truth structure sets plus 45 contoured by one observer). Execution time was measured for each method. Autosegmented structures were compared to ground truth structures using the Dice similarity coefficient, mean surface distance, Hausdorff distance, and Jaccard index. For each metric and OAR, performance was compared to the inter-observer variability using Dunn’s test with control. Methods were compared pairwise using the Steel-Dwass test for each metric pooled across all OARs. Further dosimetric analysis was performed on three high-performing autosegmentation methods (DL, IPP with RF and 4 fractions (IPP_RF_4), IPP with 1 fraction (IPP_1)), and one low-performing (PAL with STAPLE and 5 atlases (PAL_ST_5)). For five patients, delivered doses from clinical plans were recalculated on setup images with ground truth and autosegmented structure sets. Differences in maximum and mean dose to each structure between the ground truth and autosegmented structures were calculated and correlated with geometric metrics.

**Results:** DL and IPP methods performed best overall, all significantly outperforming inter-observer variability and with no significant difference between methods in pairwise comparison. PAL methods performed worst overall; most were not significantly different from the inter-observer variability or from each other. DL was the fastest method (33 seconds per case) and PAL methods the slowest (3.7 – 13.8 minutes per case). Execution time increased with number of prior fractions/atlases for IPP and PAL. For DL, IPP_1, and IPP_RF_4, the majority (95%) of dose differences were within ±250 cGy from ground truth, but outlier differences up to 785 cGy occurred. Dose differences were much higher for PAL_ST_5, with outlier differences up to 1920 cGy. Dose differences showed weak but significant correlations with all geometric metrics (R^2^ between 0.030 and 0.314).

**Conclusions:** The autosegmentation methods offering the best combination of performance and execution time are DL and IPP_1. Dose reconstruction on on-board T2-weighted MRIs is feasible with autosegmented structures with minimal dosimetric variation from ground truth, but contours should be visually inspected prior to dose reconstruction in an end-to-end dose accumulation workflow.

## Introduction

Novel magnetic resonance imaging (MRI)-linear accelerator (MR-linac) devices have enabled the clinical adoption of on-board adaptive radiation therapy (RT) [1–4] for head and neck cancers. While MR-linac systems improve soft tissue visualization and allow a new RT treatment plan to be created during every treatment fraction to better target the tumor and avoid healthy tissues, current systems do not have mechanisms for accumulating delivered dose over the entire course of RT.

Furthermore, on the 1.5T MR-linac system, the Adapt to Position workflow (virtual isocenter shift) is used for the majority of fractions in standard-fractionation head and neck RT [4], which minimizes treatment times compared to the Adapt to Shape (full adaptive replan) workflow and is appropriate when day-to-day anatomical variations are small [5]. However, this workflow reduces the time required for adaptive replanning by estimating dose on the previously segmented reference image rather than calculating dose directly on the setup image, which reflects the anatomy at the time of beam delivery. Thus, there remains an unmet need for a method to not only accumulate dose across multiple fractions but also to accurately reconstruct the delivered dose on the anatomy at the time of treatment for Adapt to Position plans.

Our approach for dose accumulation for head and neck adaptive RT on the MR-linac involves 1) autosegmenting the structures on each fraction’s setup image, 2) recalculating the delivered dose on the setup image, 3) deformably registering each setup image to a common time point (i.e. the pre-RT or post-RT anatomy), 4) deformably mapping doses to that same common time point, and 5) summing the mapped doses. In this paper, we are focusing on the first two steps to determine the optimal method for automatically segmenting the T2-weighted setup images used in our head and neck workflow [4] and to evaluate the impact of autosegmentation on dose calculation accuracy.

Various autosegmentation methods that have historically been applied to computed tomography (CT) images have shown promise on MRI [6], including deformable image registration (DIR)-based structure propagation [4,7], atlas-based autosegmentation [8–10], and deep learning [11–13]. While several studies have shown that deep learning can improve organ-at-risk (OAR) segmentation accuracy compared to atlas-based autosegmentation on CT for head and neck [14–16] and other treatment sites [17–20], to our knowledge, only a single study thus far has directly compared these methods on MRI for any treatment site [21]. As MR-guided adaptive RT becomes more accessible, evaluating these autosegmentation methods on MRI is crucial. We are also interested in leveraging images from previous MR-linac fractions to contour future fraction images for the same patient. Images from multiple prior fractions can be used as atlases in an individualized patient prior atlas-based autosegmentation approach, which has been shown to improve segmentation accuracy over deformable structure propagation from a single prior image [22].

Our aim in this paper is to evaluate the geometric accuracy of these autosegmentation methods for head and neck OARs and to test permutations with various sets of parameters, including different label fusion methods and numbers of atlases. We will also explore the feasibility of recalculating daily fraction doses on autosegmented setup images from the MR-linac and understand how differences in geometric accuracy affect the recalculated dose.

## Materials and Methods

### Patients and Informed Consent

18 head and neck cancer patients and 3 healthy volunteers were included in this study. For patients, the disease sites included 9 oropharynx (8 human papilloma virus (HPV)+ and 1 HPV-), 3 larynx, 2 nasopharynx/nasal cavity/orbit, 2 oral cavity, and 1 hypopharynx. The age range for all subjects was 22 to 81 (median: 60). All patients provided written informed consent to participate in the IRB-approved MOMENTUM observational clinical trial [23] (National Clinical Trial Identifier: NCT04075305; MD Anderson Cancer Center IRB study ID: PA18-0341), and all healthy volunteers provided written informed consent to participate in an internal IRB-approved volunteer imaging study (MD Anderson Cancer Center IRB study ID: PA14-1002).

### Images and Manual Segmentation

A total of 41 T2-weighted MRI scans were used as the primary data set in this study (5 each for 5 patients and 1 each for the remaining 16 patients/volunteers, as explained below), and an additional 45 image sets from the same patients were added for the deep learning model. All images were acquired on a 1.5 T MR-linac (Unity; Elekta AB; Stockholm, Sweden). The T2-weighted sequence is a low-resolution 2-minute scan used for setup and treatment plan reoptimization in the Unity Adapt to Position workflow, as described previously [4,5], with the following scan parameters: 3D spin echo acquisition, 1535 ms repetition time, 278 ms echo time, 0.83 mm in-plane resolution, 2 mm slice thickness, 1 mm slice gap, 400 × 400 mm^2^ field of view, 300 slices; 117 second acquisition time.

Each image in the primary data set was manually segmented by 7 individual observers (postgraduate year 4 radiation oncology residents who were specifically trained on head and neck anatomy and MRI by an experienced head and neck radiation oncologist). The following OARs were segmented: brainstem, mandible, left and right parotid glands, left and right submandibular glands, and spinal cord. Ground truth contours were generated from the 7 observer contours using a multi-label simultaneous truth and performance level estimation (STAPLE) algorithm [24] constrained to prevent overlapping contours (ADMIRE software v3.26, Elekta AB, Stockholm, Sweden). All STAPLE contours were reviewed by two additional observers (one radiologist and one radiation oncologist with at least 5 years of experience) for quality, and all were considered sufficient for further use as ground truth. These ground truth contours from this primary cohort were used as atlases/training data for the atlas-based and deep learning autosegmentation methods as well as the ground truth contours used to evaluate the autosegmentation methods. The 45 additional image sets that were added to the deep learning model were contoured by a single observer (radiologist).

### Autosegmentation Methods

A total of 20 autosegmentation methods in ADMIRE were evaluated, divided into three primary categories: 1) population atlas library (PAL), 2) individualized patient prior (IPP), and 3) deep learning (DL).

#### Population Atlas Library (PAL)

PAL is conventional multi-atlas-based autosegmentation where all the atlases (i.e. image and structure set pairs) are from different patients so that the atlas library is representative of a wide range of anatomies and can be generalized to other patients. In multi-atlas-based autosegmentation, each atlas is deformably registered to the image to be segmented, resulting in one intermediate structure set per atlas. Next, a label fusion step is performed to generate a final consensus contour from all of the intermediate contours. Three label fusion methods were used: STAPLE, patch fusion (PF), and random forest (RF) (described below). For each label fusion method, the autosegmentation was run using 5, 10, and 15 atlases, resulting in 9 total PAL-based autosegmentation methods for evaluation.

STAPLE estimates the sensitivity and specificity of each intermediate segmentation as compared to the remaining segmentations (on a structure-by-structure basis), then it weights each segmentation based on its relative performance and generates a consensus segmentation based on the weighted average [24,25]. Unlike STAPLE, which uses only the label data, PF uses the intensity values of the image to create the weights for the weighted average. Specifically, the similarity between each atlas and the image to be segmented is estimated by computing the local cross-correlation coefficient (LCC) for every voxel, and the atlas weights are determined on a voxel-by-voxel basis based on the LCC [26,27]. Finally, the RF autosegmentation method uses the atlases to train a RF model—a supervised machine learning algorithm based on decision trees—to create a binary classifier to determine whether or not each voxel in an image belongs to a given structure [28]. The RF classifier is applied only to the voxels where the intermediate segmentations mapped from each atlas do not fully agree.

#### Individualized Patient Prior (IPP)

IPP is atlas-based autosegmentation using images from the same patient as atlases. With the MR-linac, an image is acquired at every fraction, so if a small number of image sets from the first few fractions are contoured, we can leverage that data to segment images from the remaining fractions. In this study, IPP was evaluated using 1, 2, 3, and 4 prior fractions. Images from the patient’s 5^th^ fraction were used for evaluation, and the fraction(s) immediately prior were used as atlases (i.e. the 4^th^ fraction was used for 1 prior fraction, the 3^rd^ and 4^th^ were used for 2 prior fractions, etc.). When one prior fraction is used, IPP is simply a DIR-based structure propagation and requires no label fusion. When multiple prior fractions are used, IPP works similarly to PAL with the same three label fusion methods (STAPLE, PF, RF). A total of 10 IPP-based autosegmentation methods were evaluated: IPP with 1 prior fraction and IPP with 2, 3, and 4 prior fractions each using STAPLE, PF, and RF.

#### Deep Learning (DL)

The DL model used a 3D ResUNet framework [29] trained on 86 image sets (41 with ground truth STAPLE consensus contours from the 7 observers plus 45 contoured by a single observer to maximize available training data). The network structure consisted of an encoder and decoder plus five long-skip connections for a total of five levels between the encoder and decoder. A residual block was used in each level of the encoder and decoder, and down-sampling and up-sampling layers (i.e. max-pooling and up-pooling operators) were used to connect each block. The MR images were pre-processed by thresholding the lower and upper 0.25% pixel values to eliminate potential outliers, normalizing pixel values using Z-score normalization, and scaling all image values into the range of [-1, 1] using a linear transform. All algorithm modules were developed using the TensorFlow DL library with a Python and C++ interface, and the final deep learning autosegmentation model was implemented into ADMIRE. An approach similar to leave-one-out cross validation was used; since multiple image sets per patient were included for some patients, all image sets from a given patient were left out for the model evaluated on that patient.

### Geometric Evaluation and Statistical Analysis

The results of each autosegmentation method were compared against the ground truth contours using the Dice similarity coefficient (DSC) [30], mean surface distance (MSD), Hausdorff distance (HD), and Jaccard Index (JI) calculated in ADMIRE. The inter-observer variability of the seven observers was measured using pair-wise comparison of each observer’s segmentations (i.e. observer 1 vs. 2, 1 vs. 3, …, 6 vs. 7) on each image set in the primary cohort. The same four geometric indices were measured for each pair of inter-observer contours. For the spinal cord contours, since some observers did not contour the spinal cord in the entire field of view, the remainder of the spinal cord contours were cropped at the inferior-most slice for each image prior to measuring the inter-observer variability metrics. However, the consensus contours were generated from the non-cropped contours, and they were all visually inspected to ensure that the consensus contours extended through the entire field of view, which they did in every case.

The results of each autosegmentation method were compared to the inter-observer variability using Dunn’s test with control [31,32] using the inter-observer variability distribution as the control. Dunn’s test with control is a non-parametric test to compare multiple distributions to a single control distribution. Reported p-values are Bonferroni corrected p-values. The test was performed separately for each structure. Each autosegmentation method was also compared pair-wise using the Steel-Dwass test for multiple comparisons [33]. The Steel-Dwass test for multiple comparisons is a non-parametric equivalent to the Tukey all-pairs comparison with cumulative experimental error correction. The comparisons were performed pooled over all structures. All statistical analyses were performed in JMP Pro (v15.0.0, SAS Institute Inc., Cary, NC). The execution time per case of each autosegmentation method was also recorded, and the mean and standard deviation execution time were calculated.

### Dosimetric Analysis

To evaluate the impact of the differences in autosegmentation performance on the reconstructed dose, the dose from the 5^th^ Adapt to Position (ATP) fraction was recalculated on the T2-weighted image for 5 patients using the contours from 4 select autosegmentation methods and the STAPLE ground truth contours (5 total plans per patient). Four autosegmentation methods were selected for further evaluation based on their geometric performance and execution time: IPP with RF and 4 prior fractions (IPP_RF_4), DL, IPP with one prior fraction (IPP_1), and PAL with STAPLE and 5 atlases (PAL_ST_5). IPP_RF_4 performed best overall for all four geometric metrics, while DL and IPP_1 demonstrated the best tradeoff between geometric performance and execution time. PAL_ST_5 was the worst performing method across all four geometric metrics and was included to demonstrate the degree to which poor autosegmentation can affect the calculated dose.

The doses from the 5^th^ fraction (all Adapt to Position plans) were reconstructed in Monaco (Research v5.59.13; Elekta AB; Stockholm, Sweden) on the setup image set for the 5^th^ fraction using the following methodology: First, a duplicate copy of the setup image was imported for each patient with contours for the seven OARs (parotid glands, submandibular glands, mandible, spinal cord, brainstem) from each of the four autosegmentation methods and the ground truth contours. The setup image was then aligned to the reference image for the ATP plan (i.e. the planning CT) using the same isocenter shift used clinically. Next, all contours that were not included in this autosegmentation study were propagated from the reference image to the setup image using the Adapt Anatomy function in Monaco, with OAR contours registered deformably and any target volumes registered rigidly. It was verified that these contours were identical across all copies of the same image so that the only difference in the structure sets would be the seven autosegmented OARs. Bulk electron densities were assigned to each structure based on the average electron density value of each structure from the CT. Next, the beams and segments were transferred from the clinically delivered ATP plan onto the setup image by creating an Adapt to Shape plan. However, instead of reoptimizing the segments and/or fluence, the dose was recalculated directly without any modification to the original beams and segments. This workflow was repeated for each of the autosegmentation results, resulting in a unique dose distribution for each autosegmentation method and the ground truth contours per patient.

To compare the dosimetric performance of each autosegmentation method, the mean dose (D_mean_) for and maximum dose (D_max_) were calculated for each of the seven structures on each recalculated dose distribution. For both D_mean_ and D_max_, the difference between the ground truth contours and each of the four autosegmentation method contours was calculated (ΔD_mean_ and ΔD_max_).

Next, we investigated whether any of the geometric metrics (DSC, HD, MSD, JI) between each autosegmented contour and the ground truth contours are correlated with the absolute value dosimetric differences (|ΔD_mean_| and |ΔD_max_|) between the autosegmented and ground truth contours. Linear regression was performed between |ΔD_mean_| and each of the four metrics and between |ΔD_max_| and each of the four metrics. The correlation coefficient (R^2^) of each linear fit was measured, and an F-test of overall significance was performed for each fit (α=0.05).

## Results

### Execution Time and Autosegmentation Failures

The mean execution time per case is shown in Figure 1. DL was the fastest method with an execution time of 33 ± 0 s (mean ± standard deviation). For all IPP and PAL cases, execution time increased with the number of prior fractions/atlases for the same autosegmentation type and label fusion method. No clear trends were observed for the execution times of the STAPLE, RF, and PF label fusion methods when the autosegmentation type and number of prior fractions/atlases were held constant.

**Figure 1:**
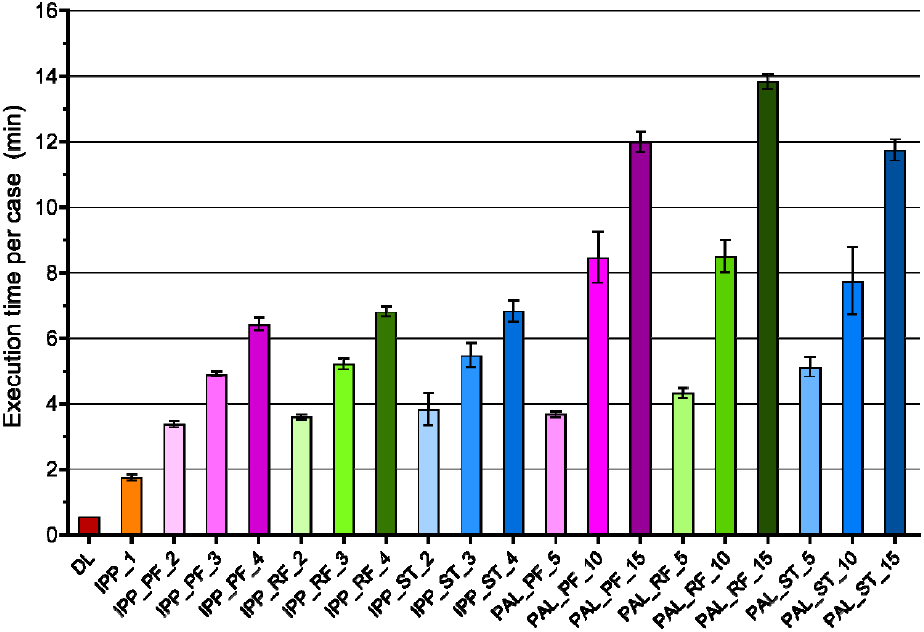
Execution time per case for each autosegmentation method. Data is represented as mean and standard deviation (error bars).

PF methods often failed in the spinal cord, meaning that no spinal cord contour could be generated despite repeated attempts. It failed in all cases for IPP_PF_2, IPP_PF_3, and IPP_PF_4. Only 4, 3, and 2 cases were successful in PAL_PF_5, PAL_PF_10, and PAL_PF_15, respectively. There were no autosegmentation failures for any other structures.

### Geometric Evaluation and Statistical Analysis

DSC and MSD values for each autosegmentation method and the pair-wise inter-observer variability analysis are shown in Figure 2. (HD and JI are shown in Appendix A.) In general, all IPP methods had a greater median DSC and JI and lower median MSD and HD compared to all PAL methods for all structures (except the DSC and JI for the mandible). PAL methods generally showed greater variability in performance among the test cases compared to IPP methods. For IPP and PAL, for a given autosegmentation type and label fusion method, no clear trends in performance were observed as the number of prior fractions or atlases increased. For PAL, PF and RF appeared to have comparable performance, but ST had poorer performance. No clear trends were observed among the label fusion methods for IPP. For most structures, DL had similar median DSC and MSD values compared to all IPP methods but had much greater variability across test cases. All IPP methods and DL had higher median DSC values, lower median MSD values, and less variability compared to the inter-observer variability for all structures. Differences between each PAL method and the inter-observer variability were less pronounced. The highest performing method overall was IPP_RF_4, which had the highest median DSC and JI and lowest median MSD and HD for all structures. The worst performing was PAL_ST_5, which demonstrated the poorest median values for nearly every metric and structure. Segmentation results for all 20 autosegmentation methods are shown on an example patient in Figure 3.

**Figure 2:**
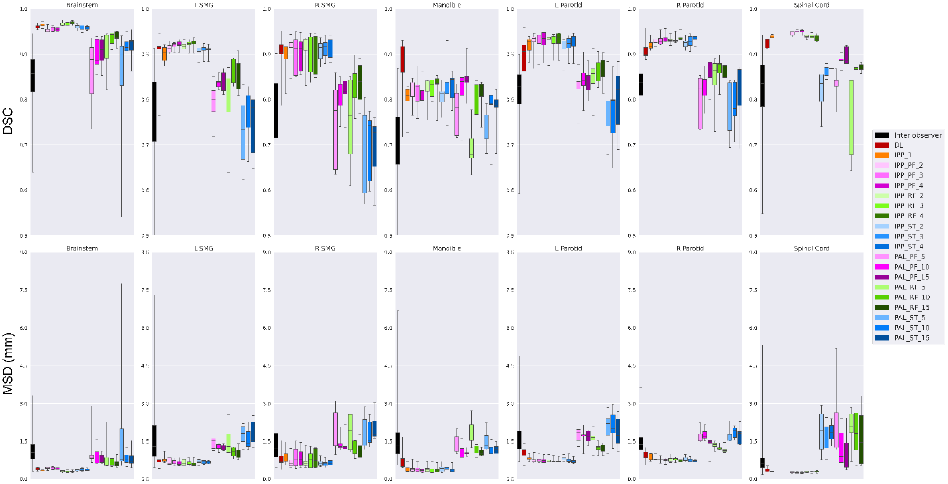
Dice similarity coefficient (DSC) and mean surface distance (MSD) in mm for the autosegmentation methods compared to ground truth contours and the pair-wise comparison of inter-observer variability. Distributions are shown as box plots, with the five horizontal bars in each distribution representing the minimum, first quartile, median, third quartile, and maximum.

**Figure 3:**
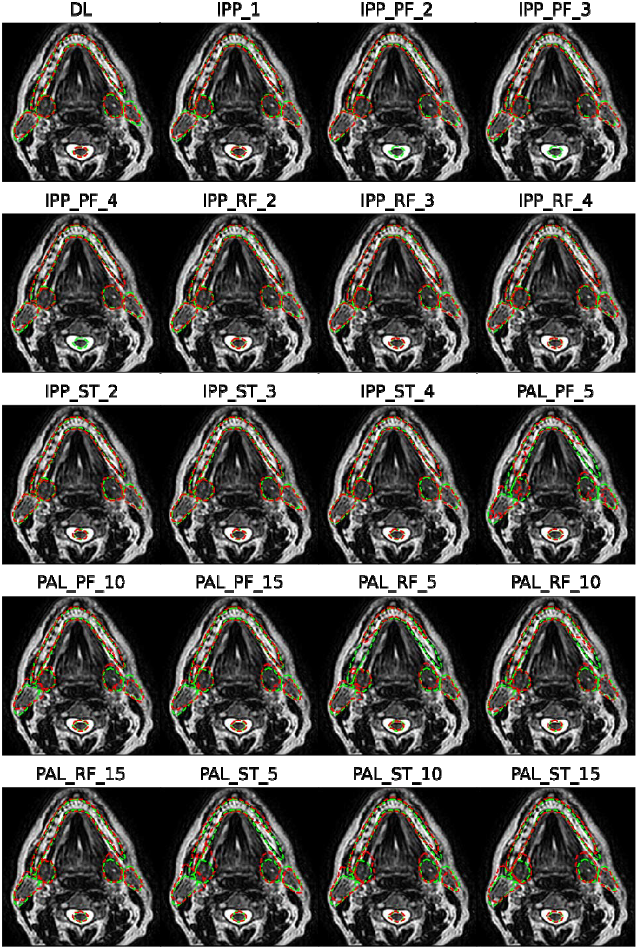
Segmentations for six of the seven OARs (excluding the brainstem since it cannot be visualized in the same plane as all the other segmentations) for one example patient. Ground truth segmentations are in green, and each set of autosegmented contours is in red. The spinal cord contour is missing in the IPP_PF methods because the segmentation failed.

Results of Dunn’s test to compare each method to the inter-observer variability are shown in Figure 4. Most IPP methods performed significantly better than the inter-observer variability for the DSC, JI, and MSD. Exceptions included the DSC and JI of IPP_1 for the mandible and MSD of all IPP_PF and IPP_RF iterations for the right submandibular gland. PAL methods were not significantly different than the inter-observer variability in most cases. There were several exceptions, mainly including the DSC and JI of the brainstem and mandible for several PAL methods with 10 and 15 atlases. Most PAL methods differed significantly from the inter-observer variability in the HD for the parotid glands. DL was significantly different than the inter-observer variability in the DSC and JI for all structures except the left submandibular gland, but it was not significantly different in the HD or MSD for all structures except the brainstem and right parotid gland.

**Figure 4:**
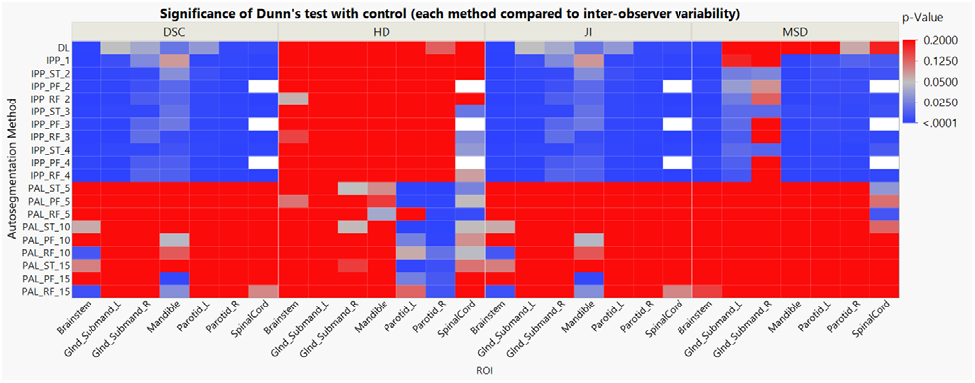
Heat map of p-values for Dunn’s test with the inter-observer variability as a control. Reported values are Bonferroni adjusted p-values. Red boxes indicate non-significant results (p>0.05), and blue boxes indicate significant results (p<0.05). White represents missing values (IPP_PF methods failed for the spinal cord for every test case). (Abbreviations: DL = deep learning, IPP = individualized patient prior, PAL = population atlas library, ST = STAPLE, PF = patch fusion, RF = random forest. 1, 2, 3, and 4 represent the number of prior fractions in IPP. 5, 10, and 15 represent the number of atlases in PAL. Glnd_Submand = submandibular gland. L = left, R = right.)

Results of the Steel-Dwass test for pair-wise comparison between each method are shown in Figure 5. None of the IPP methods or DL were significantly different from each other in any of the four metrics. Most PAL methods were not significantly different from each other with a few exceptions. Notably, PAL_ST_5 (the worst performing method overall) significantly underperformed PAL_PF_10, PAL_RF_10, PAL_PF_15, and PAL_RF_15 in most metrics. PAL_RF_15 (the best performing method overall) performed significantly better than PAL_ST_5, PAL_PF_5, and PAL_ST_10 in all metrics but HD. Most PAL methods were significantly different than all other IPP methods and DL except for PAL_RF_10 and PAL_RF_15.

**Figure 5:**
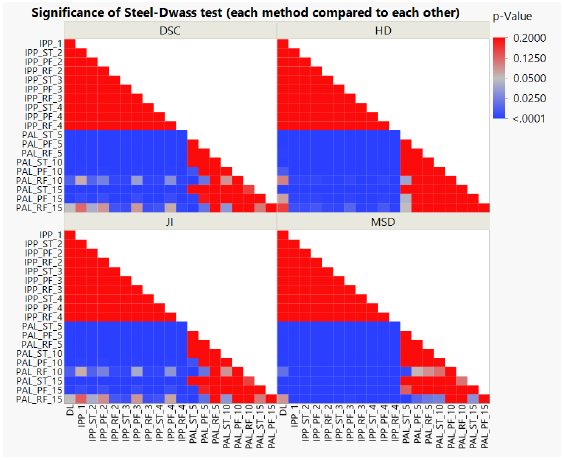
Heat map of p-values for the Steel-Dwass test for pair-wise comparison between autosegmentation methods pooled over all ROIs. Red boxes indicate non-significant results (p>0.05), and blue boxes indicate significant results (p<0.05). (Abbreviations: DL = deep learning, IPP = individualized patient prior, PAL = population atlas library, ST = STAPLE, PF = patch fusion, RF = random forest. 1, 2, 3, and 4 represent the number of prior fractions in IPP. 5, 10, and 15 represent the number of atlases in PAL.)

### Dosimetric Analysis

The dose differences (ΔD_mean_ and ΔD_max_) between plans recalculated with the ground truth contours vs. plans recalculated with the various autosegmented contours (IPP_RF_4, DL, IPP_1, and PAL_ST_5) are shown in Figure 6. For IPP_RF_4, DL, and IPP_1 (the high-performing methods geometrically), the majority (95%) of ΔD_mean_ and ΔD_max_ values across all ROIs fell within ±250 cGy. However, a few outlier dose differences occurred even for these three high-performing methods, with maximum |ΔD_mean_| and |ΔD_max_| as high as 617 cGy and 785 cGy, respectively. For these three methods, dosimetric differences greater than ±250 cGy occurred in all ROIs except the spinal cord and mandible and occurred most often in the parotid glands. Among the five cases, they occurred most often in cases 1 and 3 (treatment sites: left supraglottis and left glottis, respectively). Of the three autosegmentation method, they occurred most often for DL. For PAL_ST_5 (the poorest performing geometrically), dosimetric differences were higher overall compared to the three high-performing methods, with maximum |ΔD_mean_| and |ΔD_max_| as high as 1112 cGy and 1919 cGy, respectively. Dosimetric differences greater than ±250 cGy occurred in all ROIs except the right submandibular gland, and differences greater than ±500 cGy occurred for all ROIs except the spinal cord and right submandibular gland. The causes of major outlier dosimetric differences (greater than ±500 cGy) in all four autosegmentation methods are explored further in Appendix B. In short, they tend to occur when the superior and/or inferior boundary of an ROI falls within a steep dose gradient and is placed in the wrong slice by the autosegmentation method.

**Figure 6:**
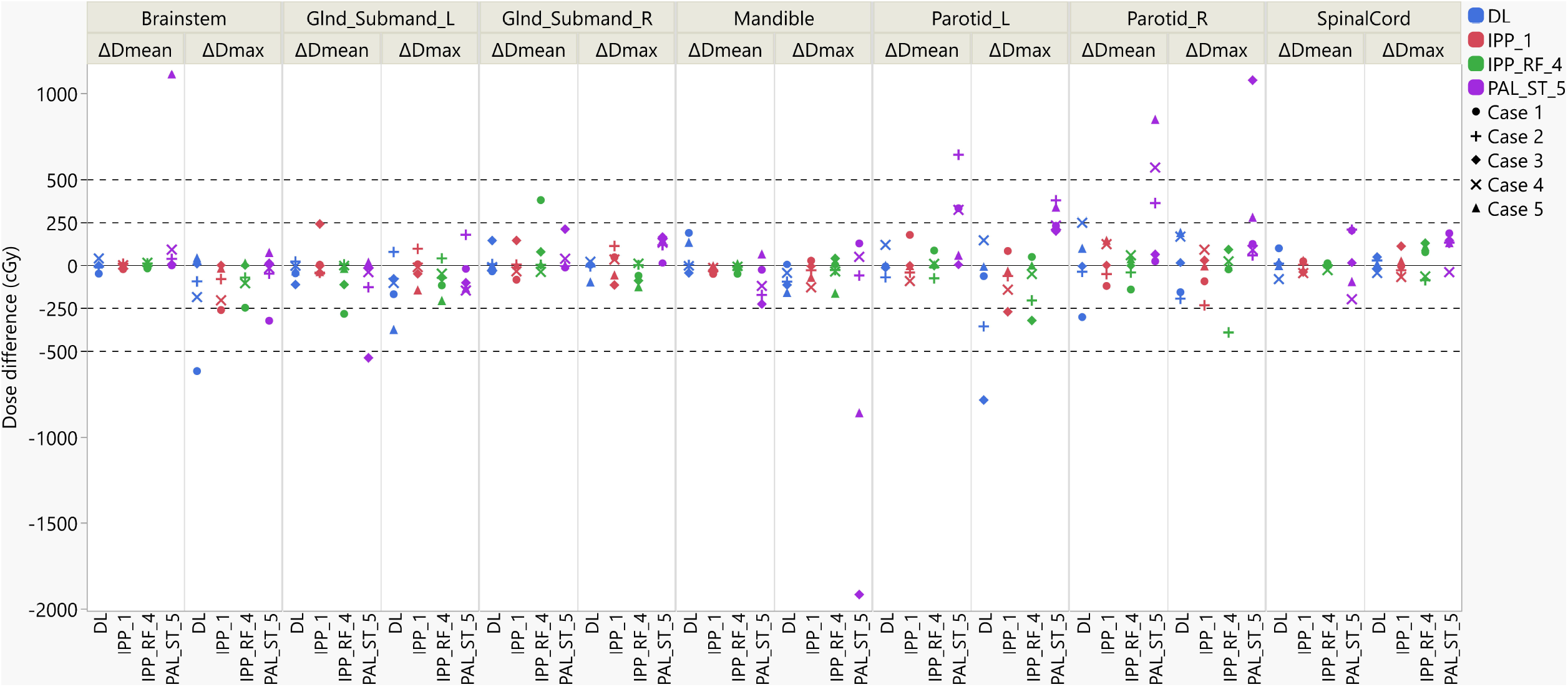
Differences (cGy) in dosimetric performance criteria (ΔD_mean_ and ΔD_max_) between plans based on ground truth contours and select autosegmentation methods for 5 patient cases. Positive/negative values mean that the dose was higher/lower in the autosegmented structure than in the ground truth structure, respectively. Treatment sites for the five patient cases are: case 1: larynx – left supraglottis; case 2: left hypopharynx; case 3: larynx – left glottis; case 4: oropharynx – right tonsil; case 5: oropharynx – right tonsil. (Abbreviations: D_mean_ = mean dose, D_max_ = maximum dose, L = left, R = right, Glnd_Submand = submandibular gland, DL = deep learning, IPP_1 = individualized patient prior with 1 prior fraction, IPP_RF_4 = individualized patient prior with 4 prior fractions and random forest, and PAL_ST_5 = population atlas library with 5 atlases and STAPLE.)

The results of the correlation between each dosimetric difference (ΔD_mean_ and ΔD_max_) and each of the four geometric metrics (DSC, MSD, HD, JI) are shown in Appendix C. Although the F-test was significant in all cases (p<0.05), correlation coefficients were poor (maximum R^2^ value 0.314). Correlations were slightly stronger for ΔD_mean_ than for ΔD_max_ (R^2^ between 0.128 and 0.314 for ΔD_mean_ and R^2^ between 0.030 and 0.096 for ΔD_max_). Of all geometric metrics, HD demonstrated the best correlation with both ΔD_mean_ and ΔD_max_.

## Discussion

In this study, we evaluated the geometric and dosimetric performance of several autosegmentation methods for the low-resolution T2-weighted MR images used for treatment positioning in the ATP workflow for head and neck cancers on the 1.5 T MR-linac [4]. In ATP, the reference plan image is aligned with the daily setup image to create a virtual isocenter shift, but the dose for the daily adaptive plan is calculated on the reference image rather than the daily setup image [5]. This workflow allows for quick plan adaptations to account for small changes in the position of the tumor at each fraction without having to segment the setup image while the patient is on the treatment table. However, if one wants to accurately accumulate the dose delivered over a patient’s entire RT course, then the delivered dose at each ATP fraction must be reconstructed on the daily setup images, which represent the anatomy at the time of beam delivery. To calculate doses on MR images, the electron density values of each imaging voxel must be estimated, which is routinely accomplished by assigning each structure on the MR a uniform electron density value, as measured either from that patient’s CT simulation image or from population reference values [34–37]. This method, commonly called *bulk density assignment*, requires the MR to be segmented. Thus, our goal in this study was to determine the optimal autosegmentation method for these daily setup MRIs and evaluate their impact on the calculation of the delivered dose.

Two recent studies have detailed a framework for daily dose reconstruction for prostate cancer RT on the MR-linac using MRI cine imaging and treatment log files [38,39]. This approach accounts for intra-fraction motion by synchronizing the motion trajectory and small units of delivered fluence based on the time stamps. While intra-fraction motion is a major consideration for many treatment sites including prostate, intra-fraction motion is minimized in head and neck cancer treatment with the use of an immobilization mask [40]. As such, we have elected to take a computationally simpler approach where we reconstruct the dose based on the static anatomy represented in the daily setup image. Our workflow is more closely aligned with previously reported studies where dose is reconstructed based on on-board setup imaging using cone beam CT [41,42] or CT-on-rails [43], with the key difference from these studies being that a unique ATP treatment plan is delivered each day rather than delivering the same plan at each fraction.

In this study, both the geometric performance and execution time of each autosegmentation method were considered in selecting the most favorable method. IPP_RF_4 (IPP with RF label fusion and four prior atlases) had the highest median DSC and JI and lowest median MSD and HD of all methods. However, it did not perform significantly differently than DL or any other IPP method in the Steel-Dwass test for any of the four metrics, and its mean execution time of 6.8 minutes per case is 12, 4, and 2 times longer than DL, IPP_1, and IPP_RF_2, respectively. DL was also not significantly different than any of the IPP methods in terms of geometric performance but was the fastest method, with an execution time of 33 seconds per case. However, the downside of DL is that it requires many more ground truth image sets for training data compared to IPP or PAL. As such, expanding our current model to include all OARs for head and neck treatment planning would require many more manual segmentations, which would be an extremely time-intensive process. IPP_1 would be a favorable alternative to DL as long as one MR image per patient is manually segmented, although any error in the initial segmentation is propagated to the next image with IPP_1. As our results have shown, increasing the number of prior fractions with IPP does not significantly improve performance but increases execution time by roughly 1.5-2 minutes for every added fraction. This study has also shown that nearly all PAL methods perform significantly worse than DL and all IPP methods and have much longer execution times. Despite these performance limitations, PAL may be a useful method for segmenting the first image for a new patient, as long as the contours are manually reviewed and edited if necessary.

To our knowledge, the present manuscript is the first to compare multi-atlas-based autosegmentation with a population atlas library (i.e. PAL), multi-atlas-based autosegmentation using individualized patient prior images (i.e. IPP), and DL-based autosegmentation. Several recent studies have investigated at least one of these methods, and many have focused on parameter optimization, as we did in this study. Van de Velde *et al*. [44] studied the optimal number of atlases using the PAL STAPLE and PF algorithms in ADMIRE (the same autosegmentation software used in the present study) to contour the brachial plexus on CT images. Looking at a range of 2-12 atlases, they found 9 to be the optimal number of atlases for both STAPLE and PF based solely on geometric metrics. Lee *et al*. [45] also tested the optimal number of atlases in head and neck CT images using MIM software, but their approach involves using a large population atlas library and selecting a small number of atlases that most closely match the patient being segmented, followed by DIR and label fusion of the intermediate results. They looked at 20, 40, 60, 80, and 100 atlases in the library and found that geometric performance generally peaked around 60 atlases, with more atlases either not improving segmentation accuracy and even degrading performance in some cases.

Schipaanboord *et al*. [46] and Van de Velde *et al*. [47] both showed that when atlas selection strategies are used to select atlases most similar to the anatomy of the patient being segmented, performance is significantly improved over using a random selection of atlases. This concept can logically be extended to support the IPP atlas-based autosegmentation method used in the current paper; using prior images from the same patient as atlases achieves the maximal similarity between the atlases and the image being segmented. Our results support this idea, with all nearly IPP methods performing significantly better than PAL methods. Zhang *et al*. [22] investigated the IPP approach with STAPLE in ADMIRE using MR-linac images of the abdomen. They demonstrated that IPP with 7 prior images, both with and without an MRI pre-processing step, improved autosegmentation quality over both rigid registration and DIR from the pre-RT MR sim. They also found that the mean DSC increased as the number of prior fractions increased for all OARs, in contrast to our results, which did not show a significant improvement with increasing the number of prior fractions in IPP. This discrepancy may be due to the fact that there is more deformation from day to day in the abdomen compared to head and neck, so the benefit of adding more prior fractions may depend on the treatment site.

In our study, we also performed further dosimetric evaluation on contours from IPP_RF_4 (the top performing method overall geometrically), DL (the fastest method and one of the high-performing methods), IPP_1 (the second fastest method and one of the high-performing methods), and PAL_ST_5 (the worst performing method overall) to better understand how contour geometry affects the dose volume histogram parameters of the recalculated doses. Large dosimetric discrepancies between ground truth and PAL_ST_5 were observed, which was expected because the geometric performance was quite poor. While dosimetric differences were smaller overall for the three high-performing methods, roughly 5% of the all measured ΔD_mean_ or ΔD_max_ data points were greater than 250 cGy and 2% were greater than 500 cGy. Appendix B provides a closer examination of outlier ΔD_mean_ or ΔD_max_ values greater than 500 cGy for the three high-performing methods and greater than 1000 cGy for PAL_ST_5. In all cases but one, the large dosimetric discrepancy was caused by disagreement between the ground truth and autosegmented contours about the slice in which the superior and/or inferior aspect of the contour begins. When the OAR boundary occurs in a high dose region, the dose differences can be substantial. Even a difference as small as 3 slices (with 1 mm slice spacing) in one example led to a ΔD_max_ value of nearly 800 cGy.

Our results correlating the geometric and dosimetric metrics in Appendix C showed a significant but weak correlation for all metrics (maximum R^2^ of 0.314). Correlations were stronger for |ΔD_max_| than for |ΔD_mean_| for each corresponding geometric metric, and they were slightly stronger for the distance-based metrics (HD and MSD) than for the volume-based metrics (DSC and JI). These results are in accordance with similar studies that have shown that geometric indices are not strongly correlated with various measures of dosimetric plan quality [48–51], highlighting the need for autosegmentation methods to be evaluated on dosimetric criteria in addition to geometric criteria [52].

There are a few limitations involved in this study. First, we acknowledge our relatively small sample size but employed cross-validation strategies where appropriate to maximize robustness of our results. Also, the physicians who manually contoured the images were postgraduate year 4 radiation oncology residents. While the inter-observer variability among this group may be greater than fully board-certified radiation oncologists, the STAPLE consensus contours used as ground truth in this study were reviewed and approved by two more experienced observers. Finally, although a variety of DL architectures and parameters could have been tested, only one DL model was used in this study due to the number of cross-validation models that had to be created and the long training time for each model. Thus, the comparison between DL and the various PAL and IPP iterations served as a pilot study of DL for this particular clinical application, and we anticipate that further refinement of our DL model and including additional data may improve performance.

In the era of adaptive RT, there is still a critical need to develop accurate, fully automated dose accumulation strategies. Our results demonstrate that several autosegmentation methods in the ADMIRE platform, particularly DL and IPP_1, are fast and highly accurate on the low-resolution T2-weighted images used for daily positioning on the MR-linac for head and neck cancers and can be used to reconstruct daily fraction doses on the anatomy at the time of treatment. Still, dosimetric accuracy may be compromised if autosegmentation errors occur in areas with high dose gradients, resulting in dosimetric discrepancies as high as 800 cGy even with these geometrically high-performing autosegmentation methods. As such, visual inspection and manual contour editing are recommended prior to dose recalculation. Alternatively, more efficient solutions have been proposed to automatically detect potential autosegmentation failures using machine learning models [53–56]. While these tools have shown promise for detecting autosegmentation errors and minimizing user intervention, more work is needed at this time to support a fully automated dose accumulation workflow. Furthermore, next steps to realize an end-to-end dose accumulation solution include autosegmentation of target volumes [57–60] and validation of methods for deformable image registration and dose mapping and summation [61,62], which were beyond the scope of this paper.

## Conclusion

Our study has demonstrated the feasibility of implementing autosegmentation for daily dose reconstruction in an off-line dose accumulation workflow for MR-guided RT for head and neck cancers. The quantitative comparison of IPP, PAL, and DL autosegmentation methods demonstrated that DL and IPP_1 both offer the best balance between accuracy and execution time. Given highly curated, accurate initial contours, IPP_1 provides the most robust results in terms of both geometric and dosimetric accuracy but takes two minutes per case. The use of additional atlases did not significantly improve performance in IPP. DL is the fastest method (30 seconds per case) and offers comparable geometric performance to all IPP methods. Unlike IPP, which will propagate any errors in the initial segmentation, DL does not require prior segmentations from the same patient but comes at the expense of somewhat random geometric and dosimetric outliers. Although DL is less robust than IPP, further parameter optimization and inclusion of more cases may improve performance. Furthermore, PAL methods demonstrated the worst overall geometric performance, largest number of dosimetric outliers, and longest execution times (4-14 minutes), so use of PAL should be reserved for cases where DL models or prior segmentations are not available. Finally, results from the dosimetric analysis show that even small errors in autosegmentation may substantially impact the calculation of the delivered dose if the region of autosegmentation failure occurs in a high dose region. Thus, contours should always be visually inspected prior to dose recalculation and manually edited when necessary to ensure robust and accurate dose accumulation.

## Data Availability

At the time of submission, data is currently being curated to upload to a public data repository.

## References

[1] Lagendijk JJW, Raaymakers BW, van Vulpen M. The Magnetic Resonance Imaging-Linac System. Semin Radiat Oncol 2014;24:207–9. https://doi.org/10.1016/j.semradonc.2014.02.009.

[2] Raaymakers BW, Jürgenliemk-Schulz IM, Bol GH, Glitzner M, Kotte ANTJ, Van Asselen B, et al. First patients treated with a 1.5 T MRI-Linac: Clinical proof of concept of a high-precision, high-field MRI guided radiotherapy treatment. Phys Med Biol 2017;62:L41–50. https://doi.org/10.1088/1361-6560/aa9517.

[3] Mutic S, Dempsey JF. The ViewRay System: Magnetic Resonance-Guided and Controlled Radiotherapy. Semin Radiat Oncol 2014;24:196–9. https://doi.org/10.1016/j.semradonc.2014.02.008.

[4] McDonald BA, Vedam S, Yang J, Wang J, Castillo P, Lee B, et al. Initial Feasibility and Clinical Implementation of Daily MR-Guided Adaptive Head and Neck Cancer Radiation Therapy on a 1.5T MR-Linac System: Prospective R-IDEAL 2a/2b Systematic Clinical Evaluation of Technical Innovation. Int J Radiat Oncol 2021;109:1606–18. https://doi.org/10.1016/j.ijrobp.2020.12.015.

[5] Winkel D, Bol GH, Kroon PS, van Asselen B, Hackett SS, Werensteijn-Honingh AM, et al. Adaptive radiotherapy: The Elekta Unity MR-linac concept. Clin Transl Radiat Oncol 2019;18:54–9. https://doi.org/10.1016/j.ctro.2019.04.001.

[6] Cardenas CE, Yang J, Anderson BM, Court LE, Brock KB. Advances in Auto-Segmentation. Semin Radiat Oncol 2019;29:185–97. https://doi.org/10.1016/j.semradonc.2019.02.001.

[7] Christiansen RL, Dysager L, Bertelsen AS, Hansen O, Brink C, Bernchou U. Accuracy of automatic deformable structure propagation for high-field MRI guided prostate radiotherapy. Radiat Oncol 2020;15:1–11. https://doi.org/10.1186/s13014-020-1482-y.

[8] Wardman K, Prestwich RJD, Gooding MJ, Speight RJ. The feasibility of atlas-based automatic segmentation of MRI for H&N radiotherapy planning. J Appl Clin Med Phys 2016;17:146–54. https://doi.org/10.1120/jacmp.v17i4.6051.

[9] Heckemann RA, Hajnal J V., Aljabar P, Rueckert D, Hammers A. Automatic anatomical brain MRI segmentation combining label propagation and decision fusion. Neuroimage 2006;33:115–26. https://doi.org/10.1016/j.neuroimage.2006.05.061.

[10] Collins DL, Pruessner JC. Towards accurate, automatic segmentation of the hippocampus and amygdala from MRI by augmenting ANIMAL with a template library and label fusion. Neuroimage 2010;52:1355–66. https://doi.org/10.1016/j.neuroimage.2010.04.193.

[11] Elguindi S, Zelefsky MJ, Jiang J, Veeraraghavan H, Deasy JO, Hunt MA, et al. Deep learning-based auto-segmentation of targets and organs-at-risk for magnetic resonance imaging only planning of prostate radiotherapy. Phys Imaging Radiat Oncol 2019;12:80– 6. https://doi.org/10.1016/j.phro.2019.11.006.

[12] Hamwood J, Schmutz B, Collins MJ, Allenby MC, Caneiro DA. OPEN A deep learning method for automatic segmentation of the bony orbit in MRI and CT images. Sci Rep 2021:1–12. https://doi.org/10.1038/s41598-021-93227-3.

[13] Savenije MHF, Maspero M, Sikkes GG, Van Der Voort Van Zyp JRN, Alexis AN, Bol GH, et al. Clinical implementation of MRI-based organs-at-risk auto-segmentation with convolutional networks for prostate radiotherapy. Radiat Oncol 2020;15:1–12. https://doi.org/10.1186/s13014-020-01528-0.

[14] van Dijk L V., Van den Bosch L, Aljabar P, Peressutti D, Both S, Steenbakkers Roel JHM, et al. Improving automatic delineation for head and neck organs at risk by Deep Learning Contouring. Radiother Oncol 2020;142:115–23. https://doi.org/10.1016/j.radonc.2019.09.022.

[15] Chen W, Li Y, Dyer BA, Feng X, Rao S, Benedict SH, et al. Deep learning vs. atlas-based models for fast auto-segmentation of the masticatory muscles on head and neck CT images. Radiat Oncol 2020;15:1–10. https://doi.org/10.1186/s13014-020-01617-0.

[16] Guo H, Wang J, Xia X, Zhong Y, Peng J, Zhang Z, et al. The dosimetric impact of deep learning-based auto-segmentation of organs at risk on nasopharyngeal and rectal cancer. Radiat Oncol 2021;16:1–14. https://doi.org/10.1186/s13014-021-01837-y.

[17] Zhang T, Yang Y, Wang J, Men K, Wang X, Deng L, et al. Comparison between atlas and convolutional neural network based automatic segmentation of multiple organs at risk in non-small cell lung cancer. Medicine (Baltimore) 2020;99:e21800. https://doi.org/10.1097/MD.0000000000021800.

[18] Ahn SH, Yeo AU, Kim KH, Kim C, Goh Y, Cho S, et al. Comparative clinical evaluation of atlas and deep-learning-based auto-segmentation of organ structures in liver cancer. Radiat Oncol 2019;14:1–13. https://doi.org/10.1186/s13014-019-1392-z.

[19] Zabel WJ, Conway JL, Gladwish A, Skliarenko J, Didiodato G, Goorts-Matthews L, et al. Clinical Evaluation of Deep Learning and Atlas-Based Auto-Contouring of Bladder and Rectum for Prostate Radiation Therapy. Pract Radiat Oncol 2021;11:e80–9. https://doi.org/10.1016/j.prro.2020.05.013.

[20] Kim H, Jung J, Kim J, Cho B, Kwak J, Jang JY, et al. Abdominal multi-organ auto-segmentation using 3D-patch-based deep convolutional neural network. Sci Rep 2020;10:1–9. https://doi.org/10.1038/s41598-020-63285-0.

[21] Kieselmann JP, Fuller CD, Gurney-Champion OJ, Oelfke U. Auto-segmentation of the parotid glands on MR images of head and neck cancer patients with deep learning strategies. MedRxiv 2020. https://doi.org/10.1101/2020.12.19.20248376.

[22] Zhang Y, Paulson E, Lim S, Hall WA, Ahunbay E, Mickevicius NJ, et al. A Patient-Specific Autosegmentation Strategy Using Multi-Input Deformable Image Registration for Magnetic Resonance Imaging–Guided Online Adaptive Radiation Therapy: A Feasibility Study. Adv Radiat Oncol 2020;5:1350–8. https://doi.org/10.1016/j.adro.2020.04.027.

[23] van Otterloo SR de M, Christodouleas JP, Blezer ELA, Akhiat H, Brown K, Choudhury A, et al. The MOMENTUM Study: An International Registry for the Evidence-Based Introduction of MR-Guided Adaptive Therapy. Front Oncol 2020;10:1328. https://doi.org/10.3389/fonc.2020.01328.

[24] Warfield SK, Zou KH, Wells WM. Simultaneous truth and performance level estimation (STAPLE): An algorithm for the validation of image segmentation. IEEE Trans Med Imaging 2004;23:903–21. https://doi.org/10.1109/TMI.2004.828354.

[25] Han X, Hoogeman MS, Levendag PC, Hibbard LS, Teguh DN, Voet P, et al. Atlas-Based Auto-segmentation of Head and Neck CT Images. In: Metaxas D, Axel L, Fichtinger G, Székely G, editors. Med. Image Comput. Comput. Interv. -- MICCAI 2008, Berlin, Heidelberg: Springer Berlin Heidelberg; 2008, p. 434–41. https://doi.org/10.1007/978-3-540-85990-1_52.

[26] Han X. A Locally Adaptive, Intensity-Based Label Fusion Method for Multi-Atlas Auto-Segmentation. Med Phys 2012;39:3960. https://doi.org/https://doi.org/10.1118/1.4736162.

[27] Coupé P, Manjón J V., Fonov V, Pruessner J, Robles M, Collins DL. Patch-based segmentation using expert priors: Application to hippocampus and ventricle segmentation. Neuroimage 2011;54:940–54. https://doi.org/10.1016/j.neuroimage.2010.09.018.

[28] Han X. Learning-Boosted Label Fusion for Multi-atlas Auto-Segmentation. In: Wu G, Zhang D, Shen D, Yan P, Suzuki K, Wang F, editors. Mach. Learn. Med. Imaging, Springer; 2013, p. 17–24. https://doi.org/10.1007/978-3-319-02267-3_3.

[29] Zhang Z, Liu Q, Wang Y. Road Extraction by Deep Residual U-Net. IEEE Geosci Remote Sens Lett 2018;15:749–53. https://doi.org/10.1109/LGRS.2018.2802944.

[30] Dice LR. Measures of the Amount of Ecologic Association Between Species. Ecology 1945;26:297–302. https://doi.org/10.2307/1932409.

[31] Dunn OJ. Multiple Comparisons Using Rank Sums. Technometrics 1964;6:241–52. https://doi.org/10.1080/00401706.1964.10490181.

[32] Juneau P. Simultaneous nonparametric inference in a one-way layout using the SAS System. Proc. PharmaSUG 2004 Annu. Meet., 2004, p. SP04.

[33] Steel RGD. A Rank Sum Test for Comparing All Pairs of Treatments. Technometrics 1960;2:197. https://doi.org/10.2307/1266545.

[34] Owrangi AM, Greer PB, Glide-Hurst CK. MRI-only treatment planning: Benefits and challenges. Phys Med Biol 2018;63:05TR01. https://doi.org/10.1088/1361-6560/aaaca4.

[35] Prior P, Chen X, Gore E, Johnstone C, Li XA. Technical Note: Is bulk electron density assignment appropriate for MRI-only based treatment planning for lung cancer. Med Phys 2017;44:3437–43. https://doi.org/10.1002/mp.12267.

[36] Kim J, Garbarino K, Schultz L, Levin K, Movsas B, Siddiqui MS, et al. Dosimetric evaluation of synthetic CT relative to bulk density assignment-based magnetic resonance-only approaches for prostate radiotherapy. Radiat Oncol 2015;10:1–9. https://doi.org/10.1186/s13014-015-0549-7.

[37] Hsu S-H, Zawisza I, O’Grady K, Peng Q, Tomé WA. Towards abdominal MRI-based treatment planning using population-based Hounsfield units for bulk density assignment. Phys Med Biol 2018;63. https://doi.org/10.1088/1361-6560/aacfb1.

[38] Menten MJ, Mohajer JK, Nilawar R, Bertholet J, Dunlop A, Pathmanathan AU, et al. Automatic reconstruction of the delivered dose of the day using MR-linac treatment log files and online MR imaging. Radiother Oncol 2020;145:88–94. https://doi.org/10.1016/j.radonc.2019.12.010.

[39] Kontaxis C, de Muinck Keizer DM, Kerkmeijer LGW, Willigenburg T, den Hartogh MD, van der Voort van Zyp JRN, et al. Delivered dose quantification in prostate radiotherapy using online 3D cine imaging and treatment log files on a combined 1.5T magnetic resonance imaging and linear accelerator system. Phys Imaging Radiat Oncol 2020;15:23–9. https://doi.org/10.1016/j.phro.2020.06.005.

[40] Bruijnen T, Stemkens B, Terhaard CHJ, Lagendijk JJW, Raaijmakers CPJ, Tijssen RHN. Intrafraction motion quantification and planning target volume margin determination of head-and-neck tumors using cine magnetic resonance imaging. Radiother Oncol 2019;130:82–8. https://doi.org/10.1016/j.radonc.2018.09.015.

[41] McCulloch MM, Lee C, Rosen BS, Kamp JD, Lockhart CM, Lee JY, et al. Predictive Models to Determine Clinically Relevant Deviations in Delivered Dose for Head and Neck Cancer. Pract Radiat Oncol 2019;9:e422–31. https://doi.org/10.1016/j.prro.2019.02.014.

[42] Veiga C, McClelland J, Moinuddin S, Lourenço A, Ricketts K, Annkah J, et al. Toward adaptive radiotherapy for head and neck patients: Feasibility study on using CT-to-CBCT deformable registration for “dose of the day” calculations. Med Phys 2014;41:031703. https://doi.org/10.1118/1.4864240.

[43] Heukelom J, Kantor ME, Mohamed ASR, Elhalawani H, Kocak-Uzel E, Lin T, et al. Differences between planned and delivered dose for head and neck cancer, and their consequences for normal tissue complication probability and treatment adaptation. Radiother Oncol 2020;142:100–6. https://doi.org/10.1016/j.radonc.2019.07.034.

[44] Van de Velde J, Wouters J, Vercauteren T, De Gersem W, Achten E, De Neve W, et al. Optimal number of atlases and label fusion for automatic multi-atlas-based brachial plexus contouring in radiotherapy treatment planning. Radiat Oncol 2016;11:1–9. https://doi.org/10.1186/s13014-015-0579-1.

[45] Lee H, Lee E, Kim N, Kim J ho, Park K, Lee H, et al. Clinical evaluation of commercial atlas-based auto-segmentation in the head and neck region. Front Oncol 2019;9:1–9. https://doi.org/10.3389/fonc.2019.00239.

[46] Schipaanboord B, Boukerroui D, Peressutti D, Van Soest J, Lustberg T, Dekker A, et al. An Evaluation of Atlas Selection Methods for Atlas-Based Automatic Segmentation in Radiotherapy Treatment Planning. IEEE Trans Med Imaging 2019;38:2654–64. https://doi.org/10.1109/TMI.2019.2907072.

[47] Van de Velde J, Wouters J, Vercauteren T, De Gersem W, Achten E, De Neve W, et al. The effect of morphometric atlas selection on multi-atlas-based automatic brachial plexus segmentation. Radiat Oncol 2015;10:1–8. https://doi.org/10.1186/s13014-015-0570-x.

[48] Bell L, Holloway L, Bruheim K, Petrič P, Kirisits C, Tanderup K, et al. Dose planning variations related to delineation variations in MRI-guided brachytherapy for locally advanced cervical cancer. Brachytherapy 2020;19:146–53. https://doi.org/10.1016/j.brachy.2020.01.002.

[49] Tsuji SY, Hwang A, Weinberg V, Yom SS, Quivey JM, Xia P. Dosimetric Evaluation of Automatic Segmentation for Adaptive IMRT for Head-and-Neck Cancer. Int J Radiat Oncol Biol Phys 2010;77:707–14. https://doi.org/10.1016/j.ijrobp.2009.06.012.

[50] Kaderka R, Gillespie EF, Mundt RC, Bryant AK, Sanudo-Thomas CB, Harrison AL, et al. Geometric and dosimetric evaluation of atlas based auto-segmentation of cardiac structures in breast cancer patients. Radiother Oncol 2019;131:215–20. https://doi.org/10.1016/j.radonc.2018.07.013.

[51] Rigaud B, Anderson BM, Yu ZH, Gobeli M, Cazoulat G, Söderberg J, et al. Automatic Segmentation Using Deep Learning to Enable Online Dose Optimization During Adaptive Radiation Therapy of Cervical Cancer. Int J Radiat Oncol Biol Phys 2021;109:1096–110. https://doi.org/10.1016/j.ijrobp.2020.10.038.

[52] Sherer M V., Lin D, Elguindi S, Duke S, Tan LT, Cacicedo J, et al. Metrics to evaluate the performance of auto-segmentation for radiation treatment planning: A critical review. Radiother Oncol 2021;160:185–91. https://doi.org/10.1016/j.radonc.2021.05.003.

[53] Rhee DJ, Cardenas CE, Elhalawani H, McCarroll R, Zhang L, Yang J, et al. Automatic detection of contouring errors using convolutional neural networks. Med Phys 2019;46:5086–97. https://doi.org/10.1002/mp.13814.

[54] McIntosh C, Svistoun I, Purdie TG. Groupwise conditional random forests for automatic shape classification and contour quality assessment in radiotherapy planning. IEEE Trans Med Imaging 2013;32:1043–57. https://doi.org/10.1109/TMI.2013.2251421.

[55] Chen HC, Tan J, Dolly S, Kavanaugh J, Anastasio MA, Low DA, et al. Automated contouring error detection based on supervised geometric attribute distribution models for radiation therapy: A general strategy. Med Phys 2015;42:1048–59. https://doi.org/10.1118/1.4906197.

[56] Beasley WJ, McWilliam A, Slevin NJ, Mackay RI, Van Herk M. An automated workflow for patient-specific quality control of contour propagation. Phys Med Biol 2016;61:8577–86. https://doi.org/10.1088/1361-6560/61/24/8577.

[57] Cardenas CE, McCarroll RE, Court LE, Elgohari BA, Elhalawani H, Fuller CD, et al. Deep Learning Algorithm for Auto-Delineation of High-Risk Oropharyngeal Clinical Target Volumes With Built-In Dice Similarity Coefficient Parameter Optimization Function. Int J Radiat Oncol Biol Phys 2018;101:468–78. https://doi.org/10.1016/j.ijrobp.2018.01.114.

[58] Cardenas CE, Anderson BM, Aristophanous M, Yang J, Rhee DJ, McCarroll RE, et al. Auto-delineation of oropharyngeal clinical target volumes using 3D convolutional neural networks. Phys Med Biol 2018;63. https://doi.org/10.1088/1361-6560/aae8a9.

[59] Cardenas CE, Beadle BM, Garden AS, Skinner HD, Yang J, Rhee DJ, et al. Generating High-Quality Lymph Node Clinical Target Volumes for Head and Neck Cancer Radiation Therapy Using a Fully Automated Deep Learning-Based Approach. Int J Radiat Oncol Biol Phys 2021;109:801–12. https://doi.org/10.1016/j.ijrobp.2020.10.005.

[60] Wahid KA, Ahmed S, He R, van Dijk L V., Teuwen J, McDonald BA, et al. Development of a High-Performance Multiparametric MRI Oropharyngeal Primary Tumor Auto-Segmentation Deep Learning Model and Investigation of Input Channel Effects: Results from a Prospective Imaging Registry. MedRXiv 2021. https://doi.org/10.1101/2021.07.27.21261114.

[61] Jaffray DA, Lindsay PE, Brock KK, Deasy JO, Tomé WA. Accurate Accumulation of Dose for Improved Understanding of Radiation Effects in Normal Tissue. Int J Radiat Oncol Biol Phys 2010;76:S135–9. https://doi.org/10.1016/j.ijrobp.2009.06.093.

[62] Chetty IJ, Rosu-Bubulac M. Deformable Registration for Dose Accumulation. Semin Radiat Oncol 2019;29:198–208. https://doi.org/10.1016/j.semradonc.2019.02.002.

