## Supplement for "Investigation of Autosegmentation Techniques on T2-Weighted MRI for Off-line Dose Reconstruction in MR-Linac Adapt to Position Workflow for Head and Neck Cancers"

**Appendix A**

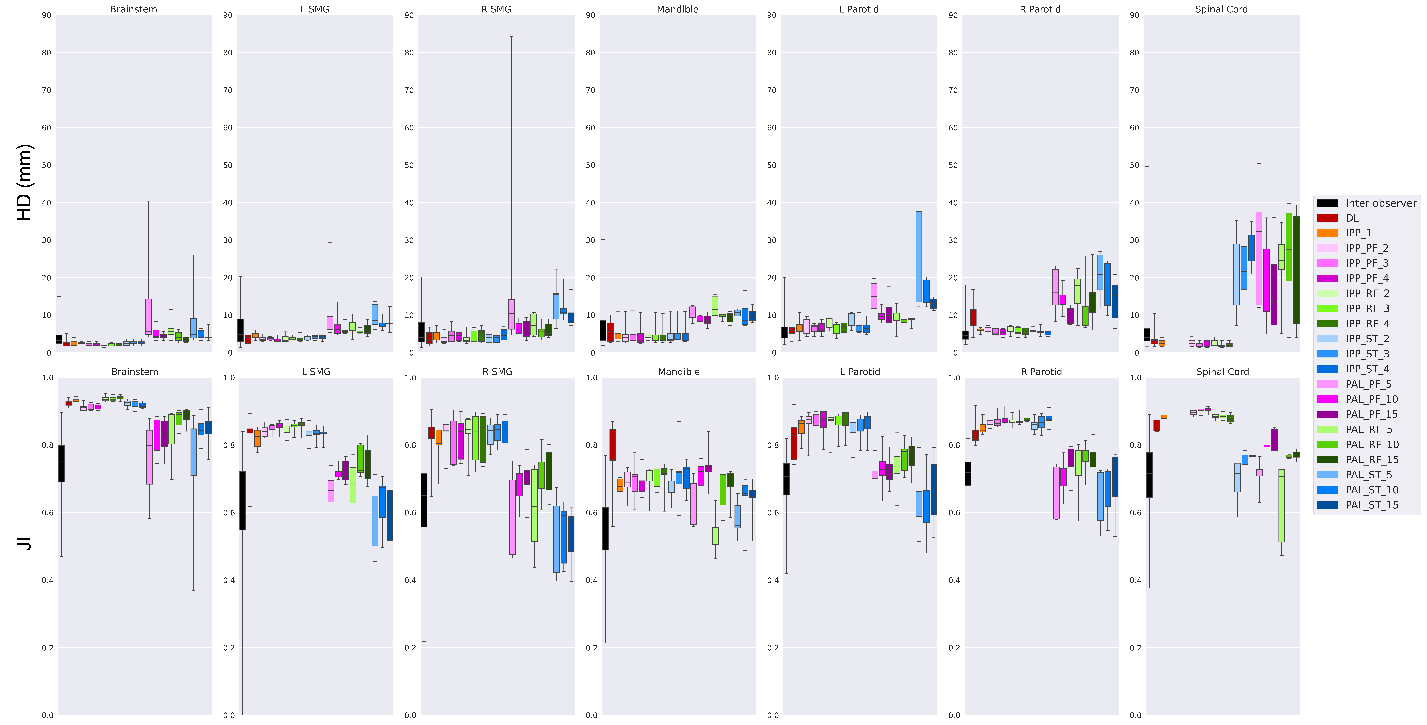

**Figure A.1:** Hausdorff distance (HD) in mm and Jaccard index (JI) for the autosegmentation methods compared to ground truth contours and the pair-wise comparison of inter-observer variability. Distributions are shown as box plots, with the five horizontal bars in each distribution representing the minimum, first quartile, median, third quartile, and maximum.

**Appendix B**

In this section, we will explore the causes of major dosimetric discrepancies between plans recalculated with the ground truth contours and the same plans recalculated with various autosegmented contours. We will look at cases where a dosimetric discrepancy was greater than 500 cGy for the three high-performing autosegmentation methods (IPP_RF_4, IPP_1, and DL) and greater than 1000 cGy for the low-performing autosegmentation method (PAL_ST_5).

Note: In all figures shown below, the spacing between slices is 1 mm.

*1. Case 1 (larynx), DL, D_max_ of Brainstem*

D_max_ of Brainstem is 3540.0 cGy with ground truth contours and 2923.0 cGy with DL contours (ΔD_max_ =

-617.0 cGy). The DSC, MSD, HD, and JI of the DL Brainstem and ground truth Brainstem contours were 0.962, 0.439 mm, 5.122 mm, and 0.927, respectively.

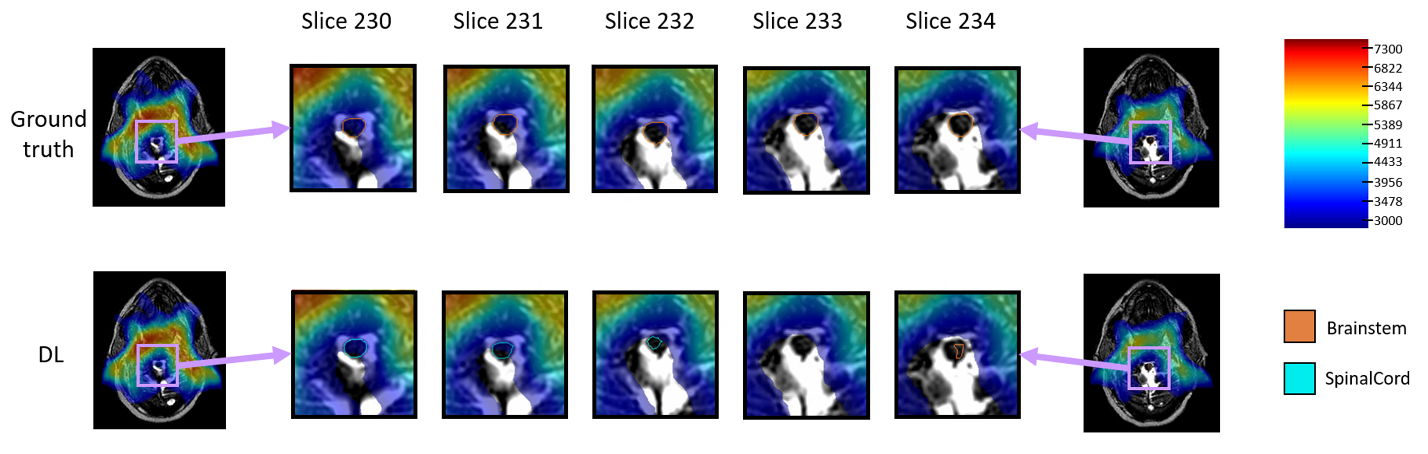

**Figure B.1:** The discrepancy in the D_max_ value of the brainstem is caused by disagreement over the slice in which the inferior aspect of the brainstem contour begins. In the ground truth segmentations, the spinal cord ends on slice 229, and the brainstem begins on slice 230. The DL model ended the spinal cord segmentation on slice 232 and began the brainstem segmentation on slice 234. The border of these two contours occurs in a steep dose gradient, resulting in an underestimation of the brainstem D_max_ with the DL contours. Nonetheless, both D_max_ values (3540 cGy for ground truth, 2923 cGy for DL) are much lower than the clinical dose constraint of D_max_ < 5400 cGy, so this difference would not change the acceptability of this plan from a clinical standpoint.

*2. Case 3 (left true vocal cord), DL, D_max_ of Parotid_L*

D_max_ of Parotd_L is 1443.0 cGy with ground truth contours and 657.8 cGy with DL contours (ΔD_max_ =

-785.2 cGy). The DSC, MSD, HD, and JI of the DL Parotid_L and ground truth Parotid_L contours were 0.862, 1.164 mm, 6.645 mm, and 0.757, respectively.

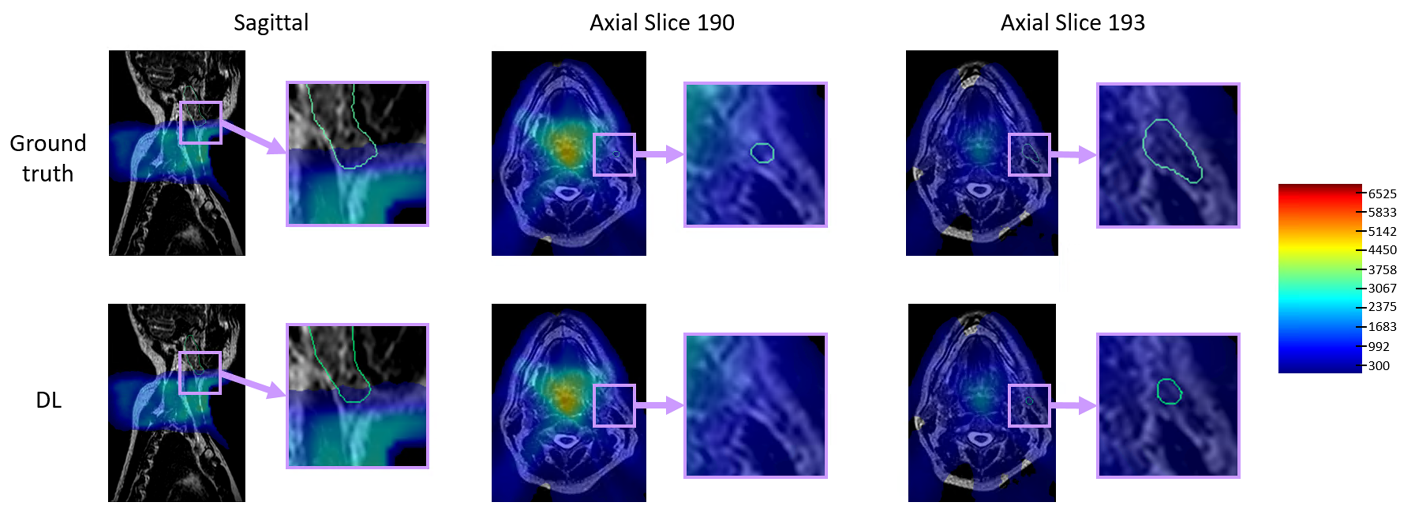

**Figure B.2:** The discrepancy in the D_max_ value of the Parotid_L is caused by disagreement over the slice in which the inferior aspect of the Parotid_L contour begins. The Parotid_L contour begins in slice 190 in the ground truth segmentation and slice 193 in the DL segmentation. In both cases, the D_max_ to Parotid_L is relatively low; in our clinical practice, we use a dose constraint of D_mean_ < 2600 cGy for the parotid glands, and the D_max_ value is lower than this threshold in both cases. However, the position of the Parotid_L contour just superior to the high dose region makes this dose discrepancy quite large, even though either recalculated dose would be considered low enough that the difference between D_max_ of 1443 cGy and 658 cGy would not change the acceptability of this plan from a clinical standpoint.

*3. Case 3 (left true vocal cord), PAL_ST_5, D_max_ of Parotid_R*

D_max_ of Parotid_R is 1057.1 cGy with ground truth contours and 2133.4 cGy with PAL_ST_5 contours (ΔD_max_ = 1076.3 cGy). The DSC, MSD, HD, and JI of the PAL_ST_5 Parotid_R and ground truth Parotid_R contours were 0.731, 2.297 mm, 26.058 mm, and 0.576, respectively.

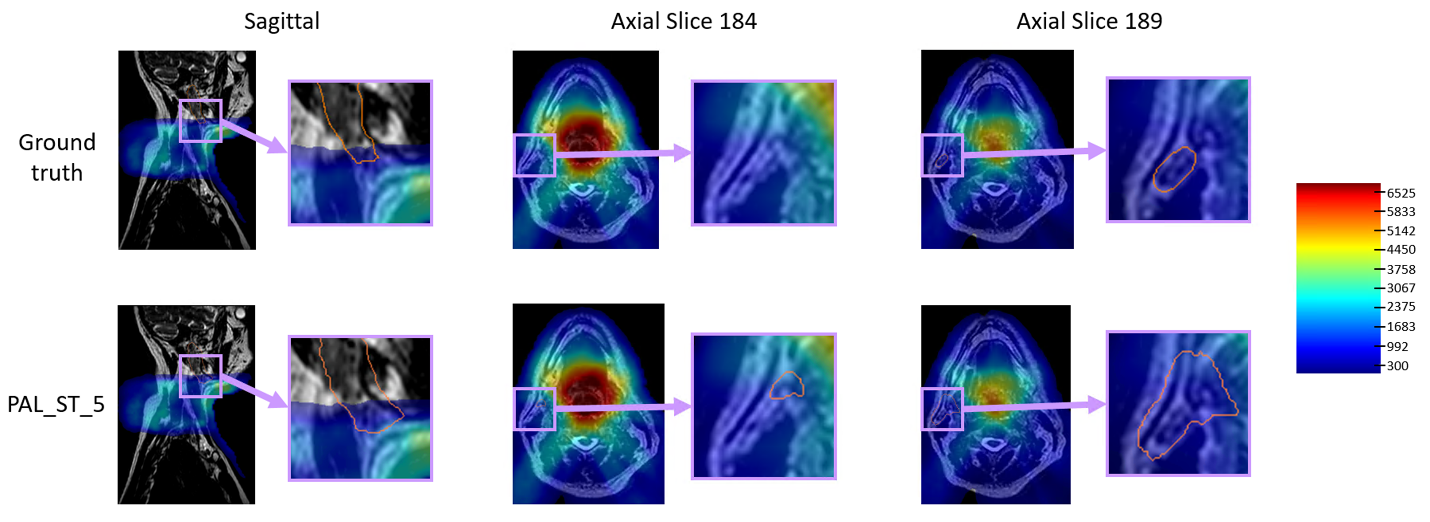

**Figure B.3:** The discrepancy in the D_max_ value of Parotid_R is caused by disagreement over the slice in which the inferior aspect of the Parotid_R contour begins. The Parotid_R contour begins in slice 189 in the ground truth segmentation and slice 184 in the PAL_ST_5 segmentation. In both cases, the D_max_ to Parotid_R is relatively low; in our clinical practice, we use a dose constraint of D_mean_ < 2600 cGy for the parotid glands, and the D_max_ value is lower than this threshold in both cases. However, the position of the Parotid_R contour just superior to the high dose region makes this dose discrepancy quite large, even though either recalculated dose would be considered low enough that the difference between D_max_ of 1057 cGy and 2133 cGy would not change the acceptability of this plan from a clinical standpoint.

*4. Case 3 (left true vocal cord), PAL_ST_5, D_max_ of Mandible*

D_max_ of Mandible is 3957.3 cGy with ground truth contours and 2038.5 cGy with PAL_ST_5 contours (ΔD_max_ = -1918.8 cGy). The DSC, MSD, HD, and JI of the PAL_ST_5 Mandible and ground truth Mandible contours were 0.681, 2.224 mm, 11.621 mm, and 0.516, respectively.

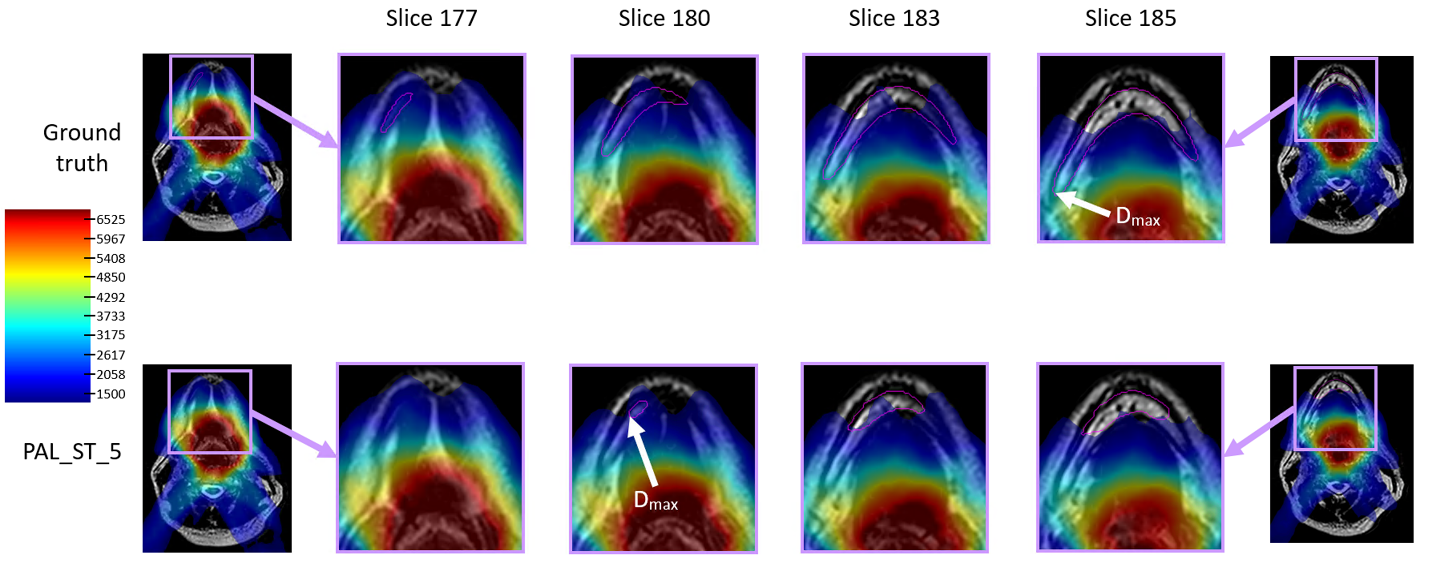

**Figure B.4:** The discrepancy in the D_max_ value of the mandible is caused by the PAL_ST_5 autosegmentation leaving out the posterior section of the mandible in the inferior slices where the mandible is adjacent to the high dose region. The point of maximum dose occurs in slice 180 for PAL_ST_5, while it occurs in slice 185 for ground truth; the PAL_ST_5 contour is missing the section of the mandible in the higher dose region in slice 185, resulting in an underestimation of the delivered dose by almost 20 Gy. There is also a disagreement over the inferior slice in which the mandible contour begins (slice 177 in the ground truth contours vs. slice 180 in the PAL_ST_5 contours). However, in this case, the inferior-most slice of the contour does not affect the calculated D_max_. Although the difference in D_max_ of nearly 20 Gy is the largest dosimetric difference found in this study, both the ground truth and PAL_ST_5 D_max_ values fall below the clinical threshold of D_max_ to the mandible < 6500 cGy and would thus not affect the acceptability of this plan from a clinical standpoint.

*5. Case 5 (right tonsil), PAL_ST_5, D_mean_ of Brainstem*

D_mean_ of Brainstem is 1116.5 cGy with ground truth contours and 2228.2 cGy with PAL_ST_5 contours (ΔD_mean_ = 1111.7 cGy). The DSC, MSD, HD, and JI of the PAL_ST_5 Brainstem and ground truth Brainstem contours were 0.541, 7.759 mm, 26.009 mm, and 0.370, respectively.

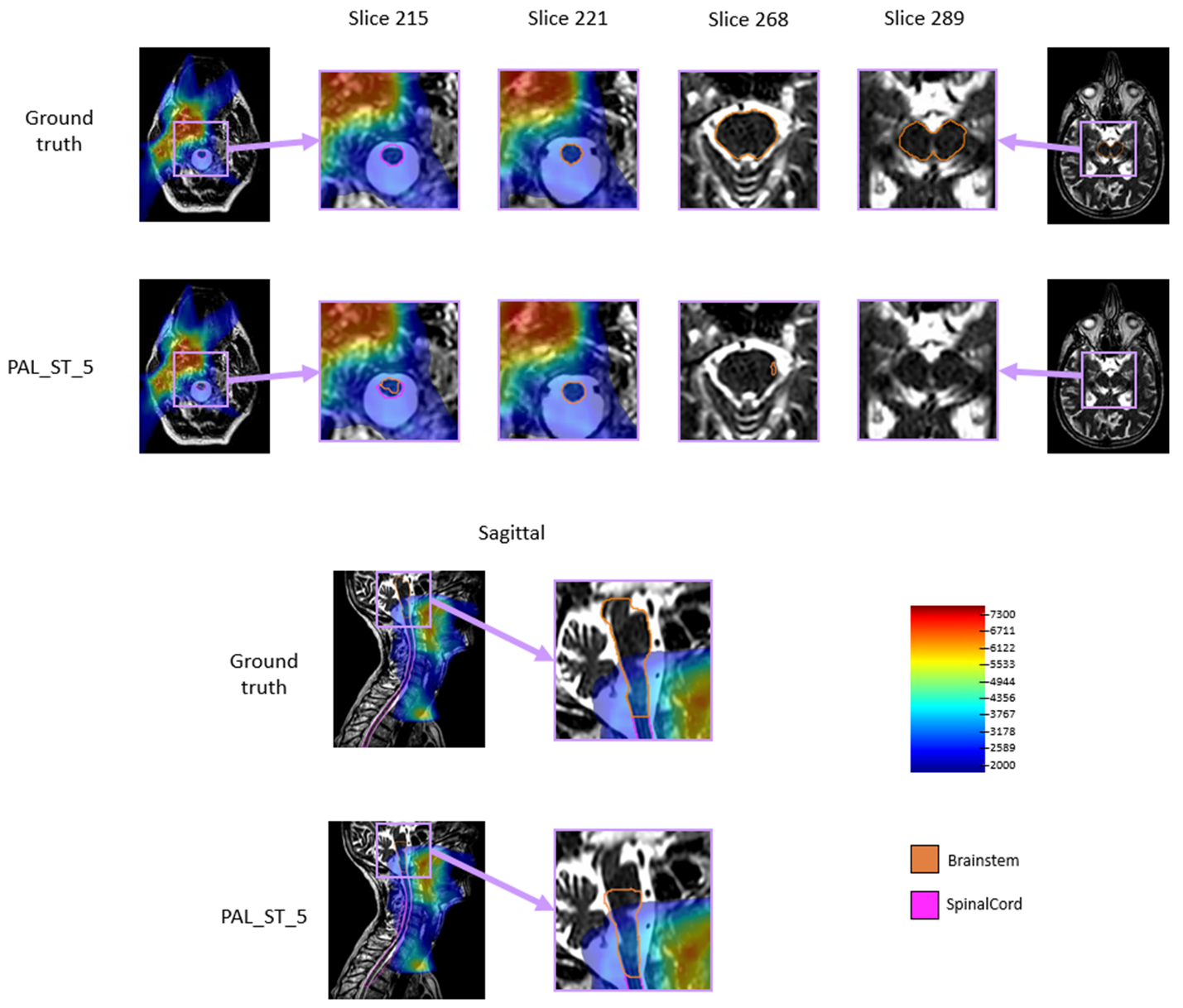

**Figure B.5:** The discrepancy in the D_mean_ value of the brainstem is caused by disagreement over the slices in which the inferior and superior aspects of the brainstem contour begin and end. The ground truth contours start in slice 221 (inferior) and end in slice 289 (superior), whereas the PAL_ST_5 contours start in slice 215 (inferior) and end in slice 268 (superior). In the PAL_ST_5 segmentations, the brainstem overlaps with the spinal cord in slices 215 and 216. Because the PAL_ST_5 brainstem contour is missing 31 slices of the superior aspect where the dose is low and adds an additional 6 slices to the inferior aspect where the dose is higher, the D_mean_ value of the brainstem is much higher for the PAL_ST_5 contours compared to the ground truth contours. Still, the clinical dose constraint of D_max_ < 5400 cGy for the brainstem is met in both cases (D_max_ = 3902 cGy for ground truth and 3977 cGy for PAL_ST_5), so this 1112 cGy discrepancy in D_mean_ would not change the acceptability of this plan from a clinical standpoint.

**Appendix C**

This section shows the correlation between the geometric and dosimetric results. For each autosegmented contour for DL, IPP_1, IPP_RF_4, and PAL_ST_5, the absolute values of the dosimetric differences from ground truth (ΔD_mean_ and ΔD_max_) were plotted against the geometric comparisons to ground truth (DSC, MSD, HD, JI). Linear regression was performed for each plot. Two outlier values were excluded (|ΔD_max_| of 1919 cGy and |ΔD_mean_| of 1112 cGy) because they were so far from the remaining points that they skewed the linear fit. The correlation coefficient (R^2^) of each linear fit was measured, and an F-test of overall significance was performed for each fit (α=0.05).

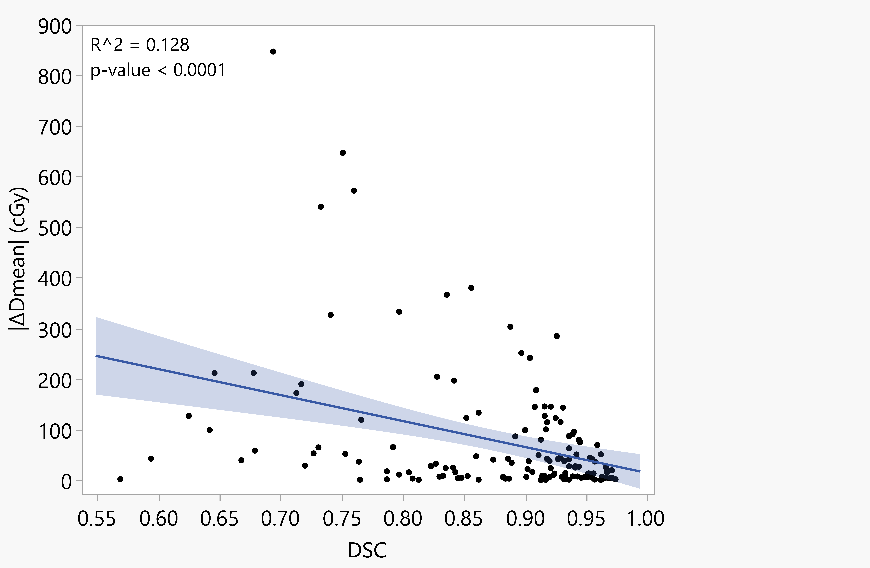

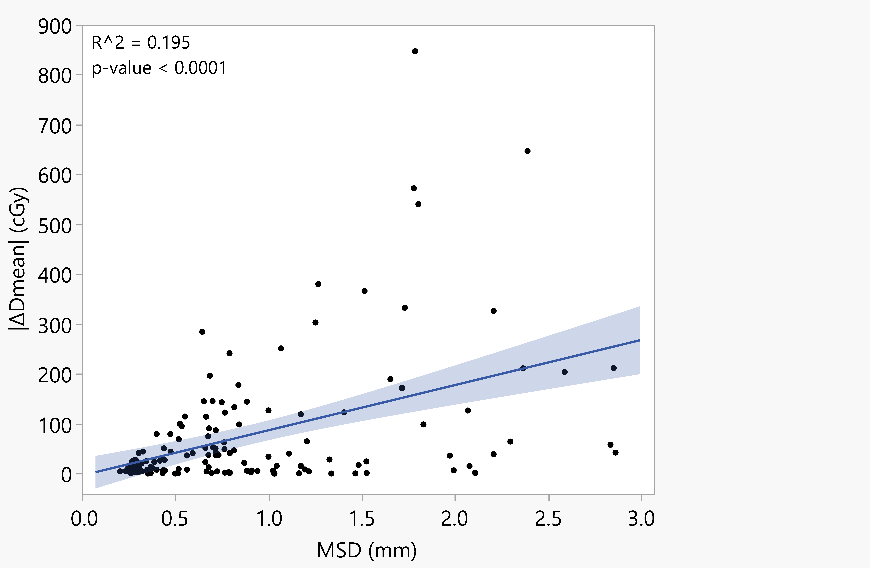

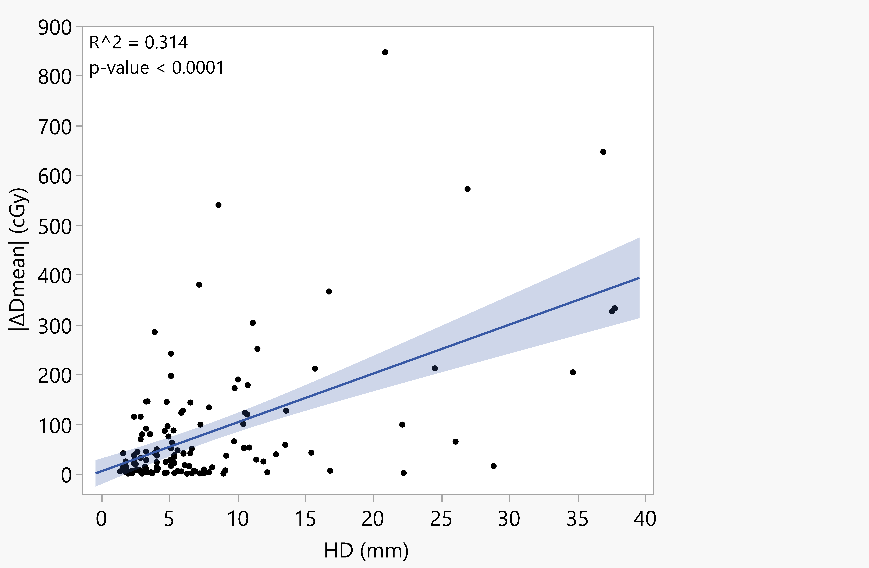

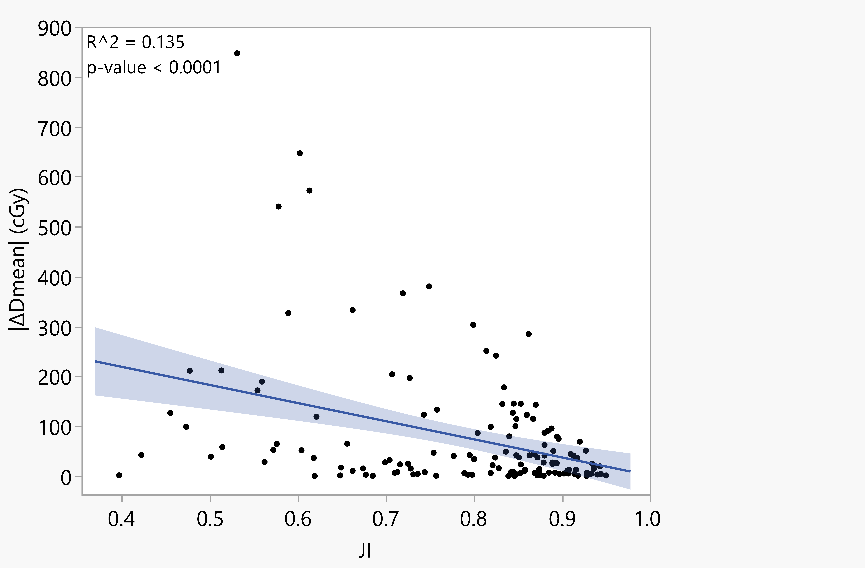

**Figure C.1:** Correlation between |ΔD_mean_| and each geometric metric. The light blue region around the line of fit shows the 95% confidence region of the fit. The correlation coefficient (R^2) and p-value of the F-test are shown in the upper left corner of each plot.

**Figure C.2:** Correlation between the absolute value of |ΔD_max_| and each geometric metric. The light blue region around the line of fit shows the 95% confidence region of the fit. The correlation coefficient (R^2) and p-value of the F-test are shown in the upper left corner of each plot.

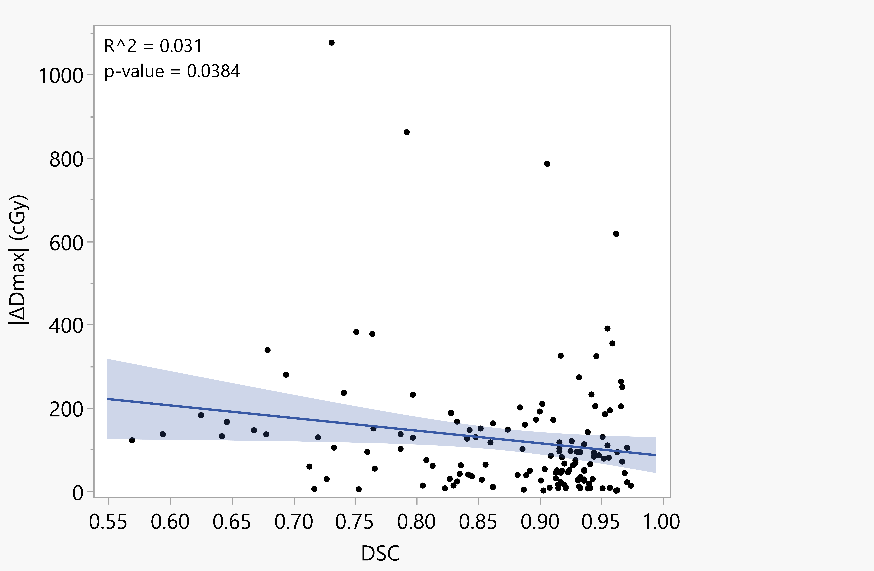

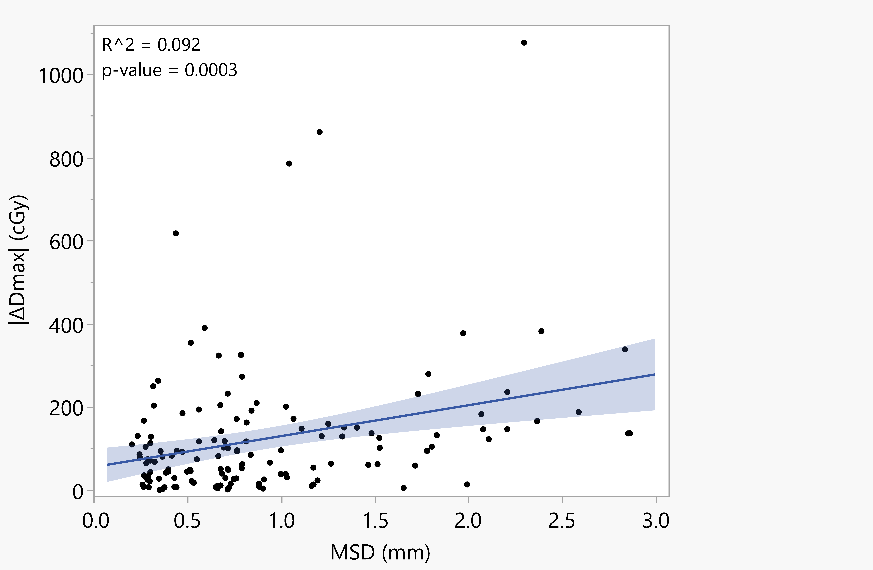

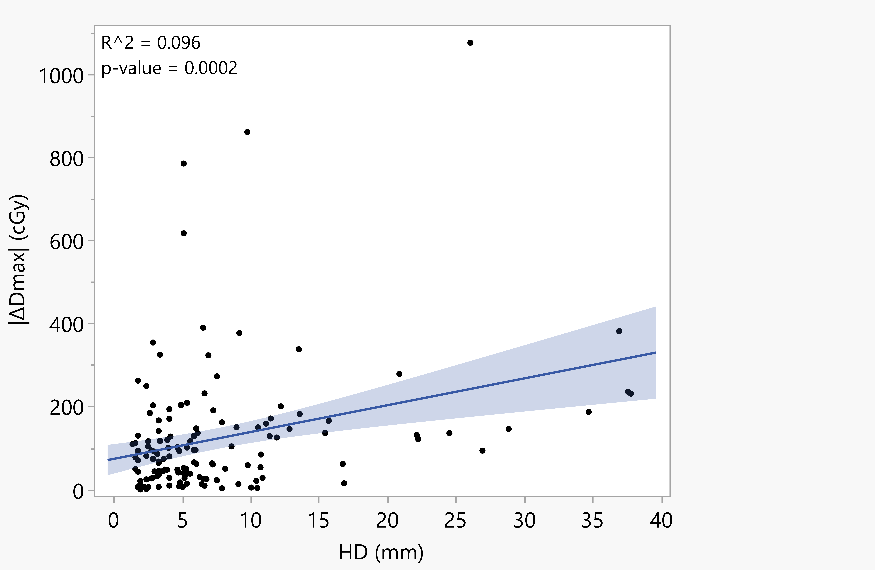

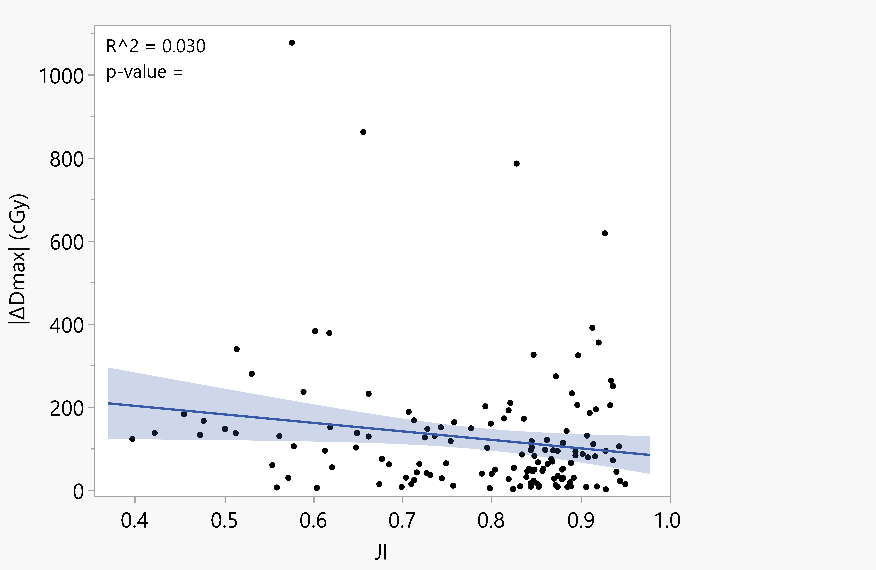

0.0422
